# Comparative Study - The Impact and Profile of COVID-19 Patients Who Are Indicated for Neuroimaging: Vascular Phenomena Are Been Found in the Brain and Olfactory Bulbs

**DOI:** 10.1101/2020.12.28.20248957

**Authors:** Maria de Fatima Viana Vasco Aragao, Mariana de Carvalho Leal, Ocelio Queiroga Cartaxo Filho, Tatiana Moreira Fonseca, Lucas Vasco Aragao, Maria Regina Vendas Carneiro Leao, Marcelo Andrade Valenca, Pedro Henrique Pereira de Andrade, Joao Pedro Vasco Aragao, Silvio da Silva Caldas Neto, Marcelo Moraes Valenca

## Abstract

**Objective:** To verify the impact and findings of the COVID-19 patients’ group that underwent brain scans in comparison to the group which only chest CT was performed.

**Method:** 876 suspected COVID-19 patients and a subsample of 232 cases with confirmed COVID-19 who underwent brain CT/MRI scan (n=35) or only chest CT (n=197) in two radiology departments, were evaluated.

**Results:** 5.59% of all suspected COVID-19 patients found had brain scans and 98.74% chest CT. There was a statistically significant difference with associations regarding the COVID-19 brain scan group for: admission to ICU, greater severity of lung injuries, the use of mechanical ventilator, seizure, sepsis, and stroke and statistical tendency for chronic renal failure and systemic arterial hypertension. 40.0% of COVID-19 patients from the brain scan group were abnormal on brain CT and/or brain MRI. 22.9% cases with any kind of bleeding or microbleeding, 8.6% with restricted diffusion lesions. One ischemic stroke case was associated with irregularity at M1 segment of the right middle cerebral artery. There was a case of left facial nerve palsy with enhancement of left geniculate ganglia. An analyse of the olfactory bulbs was possible in 12 brain MRIs and 100% had enhancement and/or microbleeding. There was no statistical difference regarding death (9.1% *versus* 5.2%).

**In conclusion:** the COVID-19 patients group on which brain CT and/or MRI needed to be performed was statically associated with the more severe COVID-19 disease, an indication to ICU, a more severe form of lung disease, use of mechanical ventilator, seizure, sepsis and stroke. Less than half of patients had abnormal brain imaging scans with all of them showing vascular brain injury lesion, being more frequently microbleeding or bleeding, followed by restricted diffusion lesions. All the olfactory bulbs evaluated showed injury by vascular phenomenon, probably methahemoglobine by microbleeding or microthrombus and/or abnormal enhancement

## Introduction

Patients with COVID-19 may develop neurological symptomatology with consequent repercussion on imaging examinations. Different abnormalities in both peripherical and central nervous systems were reported in patients with COVID-19, such as vessel wall enhancement and/or focal cerebral arteriopathy;^1^ acute ischemic infarct;^1, 2^ hemorrhage;^2, 3^ acute hemorrhagic necrotizing encephalopathy;^4^ cerebral venous thrombosis;^5, 6^ diffuse leukoencephalopathy^7^ with microhemorrhage;^8, 9^ PRES-like with microvessel enhancement;^10^ splenium of corpus callosum restricted diffusion lesion;^11^ edema^12, 13^ and enhancement and microbleeds of olfactory bulbs;^14^ as well as bilateral facial palsy and enhancement of facial nerves^15^.

Based on the principle that patients with neurological complaints, and possible intracranial lesions as attributable causes, are evaluated with brain Magnetic Resonance Imaging (MRI) or computed tomography (CT), we decided to assess two groups of patients comparatively: A group of patients with COVID-19 assessed by neuroimaging as compared to a group of COVID-19 patients who underwent only chest CT, without any image evaluation of the brain.

Thus, the purpose of this study was to verify the frequency, the clinical profile and the radiology findings of the COVID-19 patients “group that underwent brain imaging scans” in comparison to the “group of patients on which only chest CT was performed” in two radiology departments.

## Method

The Institutional Review Board approved this retrospective study of the Ethics Committee. Informed consent was waived.

A survey of the cases was carried out to identify the patients with confirmed COVID-19 who underwent chest CT scan and/or brain MRI/CT in two radiology departments.

876 clinical-suspected or confirmed COVID-19 cases were collected, being: (a) 261 patients from Radiology Department “A”, from March 28, 2020 to September 3, 2020, (11 patients underwent brain MRI and/or CT and 255 patients chest CT); and (b) 615 patients from Radiology Department “B”, from April 1, 2020 to May 24, 2020, (38 patients underwent brain MRI and/or CT and 610 patients underwent chest CT).

We used a convenience subsample of 232 cases that we were able to collect the clinical data from medical records in time to analyze and write this study which: (a) had laboratory confirmation of COVID-19, (b) excluded patients under 18 year old and with co-infection of another viral infection.

The Institution’s radiologists initially analyzed all chest and brain scans. Atypical pneumonia CT patterns for COVID-19 were excluded. The lung opacities qualitative scoring of COVID-19 (virus pneumonia) patterns was performed independently by the Institution’s radiologists: (a) less than 25%, (b) between 25% and 50% or (c) equal to or more than 50%. Our study considered only the evaluation of the Institution’s radiologists of chest CT reports.

Subsequently, all images of brain CT and MRI were reviewed independently by two radiologists and neuroradiologists who were certified by the Ministry of Education and Culture with 30 and 18 years of experience, respectively. In cases where there was disagreement, results were resolved by consensus.

The clinical variables data, retrospectively collected from the medical records, were: a) sex; b) age; c) the percentual extent of pulmonary lesions found in the chest CT; laboratory SARS-CoV2 infection confirmation; abnormalities found in brain CT or MRI, d) presence of cough, headache, anosmia, ageusia, dyspnoea, systemic arterial hypertension, dyslipidemia, obesity, renal failure, use of mechanical ventilation, and a neurological complaint. The origins of patients were also assessed, whether they were admitted to the Emergency Room, hospitalized or admitted to the ICU. It was also assessed whether a patient had progressed to death before the assessment date.

An analise of the olfactory bulbs was possible in 12 brain MRIs because 12 had at least a sequence of coronal thin slices pre- and/or post**-**contrast fat supressed T1WI and 1 patient had pre- and post-contrast 3D SPGR T1WI and also 3D FLAIR.

The intensity of olfactory bulbs was defined as normal when the bulbs have the same cortex intensity, as typically seen in healthy controls. Abnormal olfactory bulb intensity is when the bulbs are more hyperintense than the cortex on T1WI and STIR.

After gadolinium injection on T1WI, enhancement of the olfactory bulbs is defined when they become more hyperintense in comparison with their intensity on pre-gadolinium T1WI. However, when there is only the post-gadolinium T1WI and the bulb is more hyperintense than the normal cortex, this represents olfactory bulb intensity abnormality and maybe an enhancement or microbleeding (methaemoglobin), as interpreted in the present study. Microbleeding (methaemoglobin) in the olfactory bulb is considered when there is hyperintense olfactory bulb, compared with the normal cortex or the normal contralateral bulb, on pre-gadolinium fat supression TIWI.

### Statistical Analysis

Softwares SPSS 13.0 (Statistical Package for the Social Sciences) for Windows and Excel 2010 were used; all tests were applied with 95% confidence.

The results are presented in table form with their respective absolute and relative frequencies. The numerical variables are represented by measures of central tendency and measures of dispersion.

To verify the existence of an association: Chi-square Test and Fisher’s Exact Test for categorical variables were applied.

## Results

Only 49/876 (5.59%) of all suspected or confirmed COVID-19 patients had brain scans performed in both radiology departments for investigation of COVID-19 neurological complications.

The chest CT was much more requested for pulmonary complications in COVID-19 patients being 865/876 (98,74%) in both radiology departments.

In the subsample of 232 confirmed COVID-19 patients, 35 had undergone brain scans. Thirty of these 35 had chest CT performed as well. It was not taken into account whether a patient repeated the brain CT and/or MRI or the chest CT.

Thus on Table 1, the profile of the group of all the 35 confirmed COVID-19 patients that underwent brain imaging scans was compared to the convenience control group of 197 cases who only underwent chest CT (without brain scans). There were associations regarding the COVID-19 group which underwent brain imaging scans for origin within the hospital (more frequent in ICU patients), greater severity of lung injuries, the use of mechanic ventilators, complaints of dyspnea (less frequent), seizures, sepsis and ischemic stroke. There was statistical tendency for chronic kidney failure.

**Table 1.** Profile of adult patients with COVID-19 who underwent imaging scans. Comparison of the group that underwent CT and MRI of the brain with a control group that did not, but that underwent chest CT.

| Variables | CT and/or MRI of the Brain |  | p-value |
| --- | --- | --- | --- |
|  | Yes<br>n (%) | No<br>n (%) |  |
| Sex (Masculine) | 20/35 (57.1) | 115/197 (58.4) | 0.892 * |
| **Chest CT (Extension of pulmonary involvement)** |  |  |  |
| Normal | 3/31 (9.7) | 0/197 (0.0) | < **0.001** ** |
| Typical (T) < 25% | 13/31 (41.9) | 121/197 (61.4) |  |
| T between 25 - 50% | 5/31 (16.1) | 61/197 (31.0) |  |
| T >50% | 9/31 (29.0) | 14/197 (7.1) |  |
| Between 25% - 50% with pleural effusion | 0/31 (0.0) | 1/197 (0.5) |  |
| > 50% with pleural effusion | 1/31 (3.2) | 0/197 (0.0) |  |
| **Origin** |  |  |  |
| Internal | 14/33 (42.4) | 103/197 (52.3) | < **0.001** ** |
| Intensive care unit | 11/33 (33.3) | 5/197 (2.5) |  |
| Urgency | 4/33 (12.1) | 64/197 (32.5) |  |
| External | 4/33 (12.1) | 25/197 (12.7) |  |
| Systemic arterial hypertension | 18/34 (52.9) | 67/186 (36.0) | 0.062 * |
| Diabetes melitus | 10/32 (31.3) | 44/184 (23.9) | 0.376 * |
| Dyslipidemia | 2/27 (7.4) | 17/185 (9.2) | 1.000 ** |
| Asthma/COPD | 2/29 (6.9) | 25/173 (14.5) | 0.382 ** |
| Chronic kidney failure | 4/30 (13.3) | 7/182 (3.8) | 0.053 ** |
| Fever | 24/29 (82.8) | 151/188 (80.3) | 0.757 * |
| Cough | 22/29 (75.9) | 159/189 (84.1) | 0.289 ** |
| Dyspnea | 18/30 (60.0) | 151/190 (79.5) | **0.019** * |
| O<sub>2</sub> catheter | 14/29 (48.3) | 78/189 (41.3) | 0.477 * |
| Mechanic ventilatory assistance | 10/29 (34.5) | 19/190 (10.0) | **0.001** ** |
| Headache | 12/28 (42.9) | 78/188 (41.5) | 0.891 * |
| Anosmia | 7/11 (63.6) | 50/66 (75.8) | 0.462 ** |
| Disgeusia | 5/9 (55.6) | 32/43 (74.4) | 0.419 ** |
| Convulsion | 8/30 (26.7) | 2/190 (1.1) | < **0.001** ** |
| Gastric symptoms | 7/26 (26.9) | 61/190 (32.1) | 0.594 * |
| Sepsis | 7/9 (24.1) | 12/189 (6.3) | **0.006** ** |
| Obesity (qualitative) | 5/29 (17.2) | 20/186 (10.8) | 0.347 ** |
| Death | 3/33 (9.1) | 10/192 (5.2) | 0.413 ** |
| Obesity (BMI ≥ 30) | 2/9 (22.2) | 35/78 (44.9) | 0.289 ** |
| Stroke | 5/32 (15.6) | 1/171 (0.6) | < **0.001** ** |
|  | **Mean ± DP** | **Mean ± DP** |  |
| Age (years) | 54.3 ± 18.1 | 49.6 ± 13.7 | 0.163 *** |
| Body mass index | 27.1 ± 6.3 | 29.6 ± 5.2 | 0.179 *** |
|  | **Median (Q1; Q3)** | **Median (Q1; Q3)** |  |
| Interval of symptom onset at admission | 6.0 (3.7; 7.7) | 7.0 (5.0; 8.0) | 0.497 ¥ |
| Length of stay | 8.5 (5.0; 23.5) | 8.0 (5.0; 14.0) | 0.630 ¥ |

(\*) Chi-Squared, (\*\*) Fisher's Exat Test, (\*\*\*) T Test, (¥) Mann-Whitney Test
COPD, chronic obstructive pulmonary disease

No statistical differences were found between the two groups concerning fever, cough, headache, anosmia, ageusia, gastric symptoms, obesity, systemic arterial hypertension, diabetes mellitus (DM), dyslipidemia, asthma, chronic obstructive pulmonary disease and death.

Table 2 shows the individual data of demografic, clinical and radiological caracteristics of the 35 adult patients with COVID-19 on whom brain MRI and/or CT scans were performed because of clinical neurological complication. Among those 35 patients, 21 cases (60.0%) were normal and 14 (40.0%) were abnormal on brain CT and/or brain MRI (Figures 1-7), being:

**Table 2.**
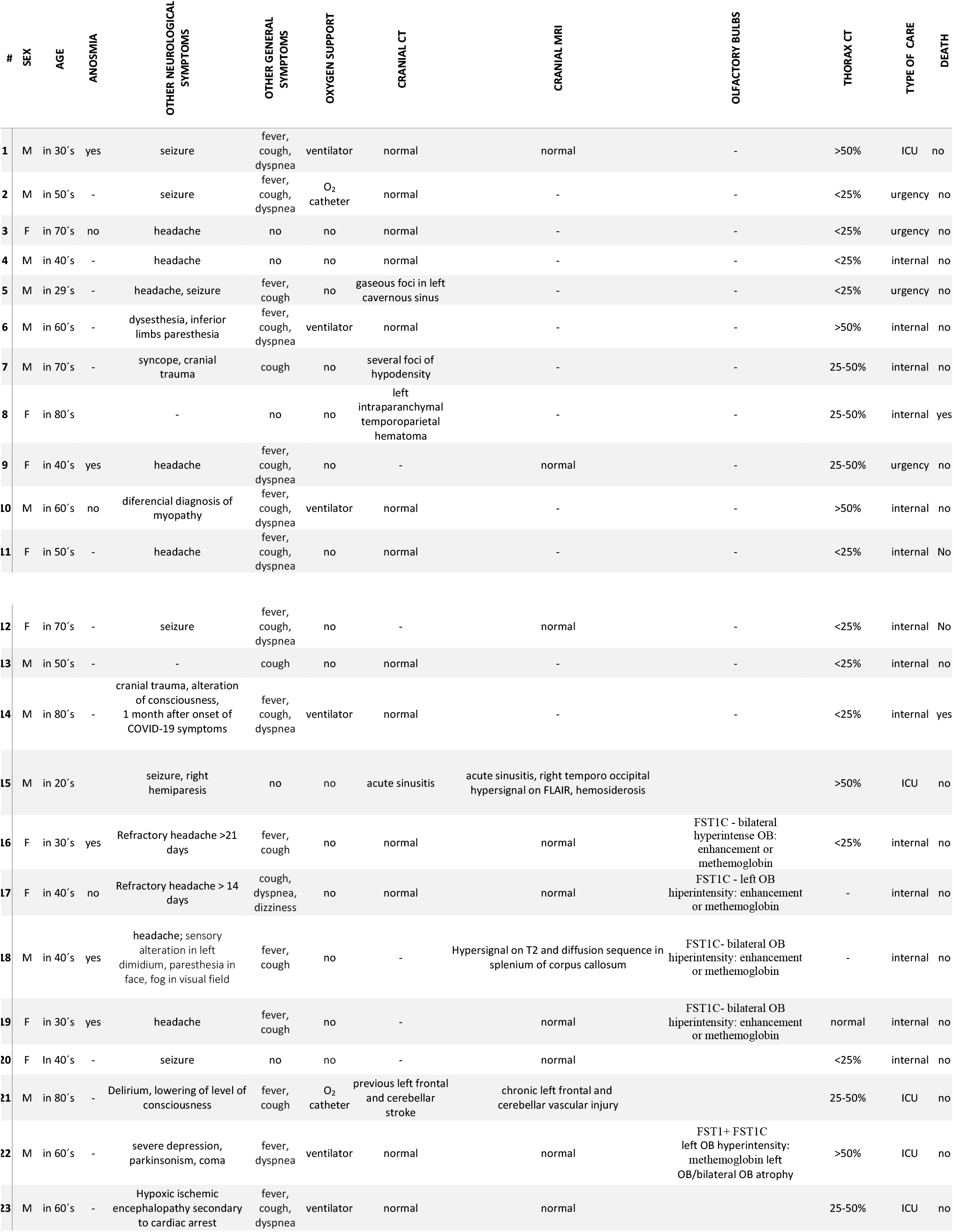
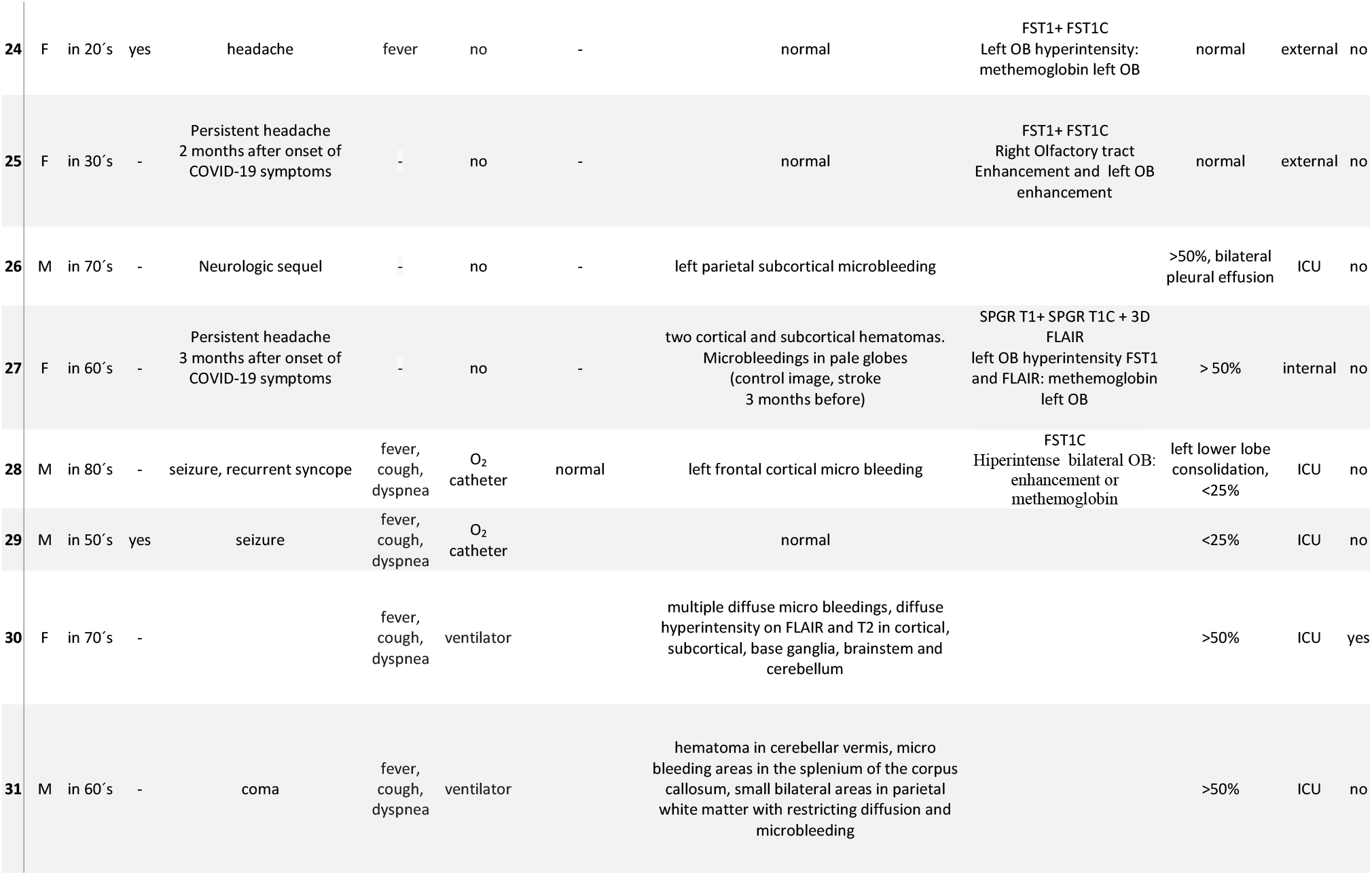
Characteristics of COVID-19 patients undergoing brain imaging (CT/MRI).

**Figure 1.**
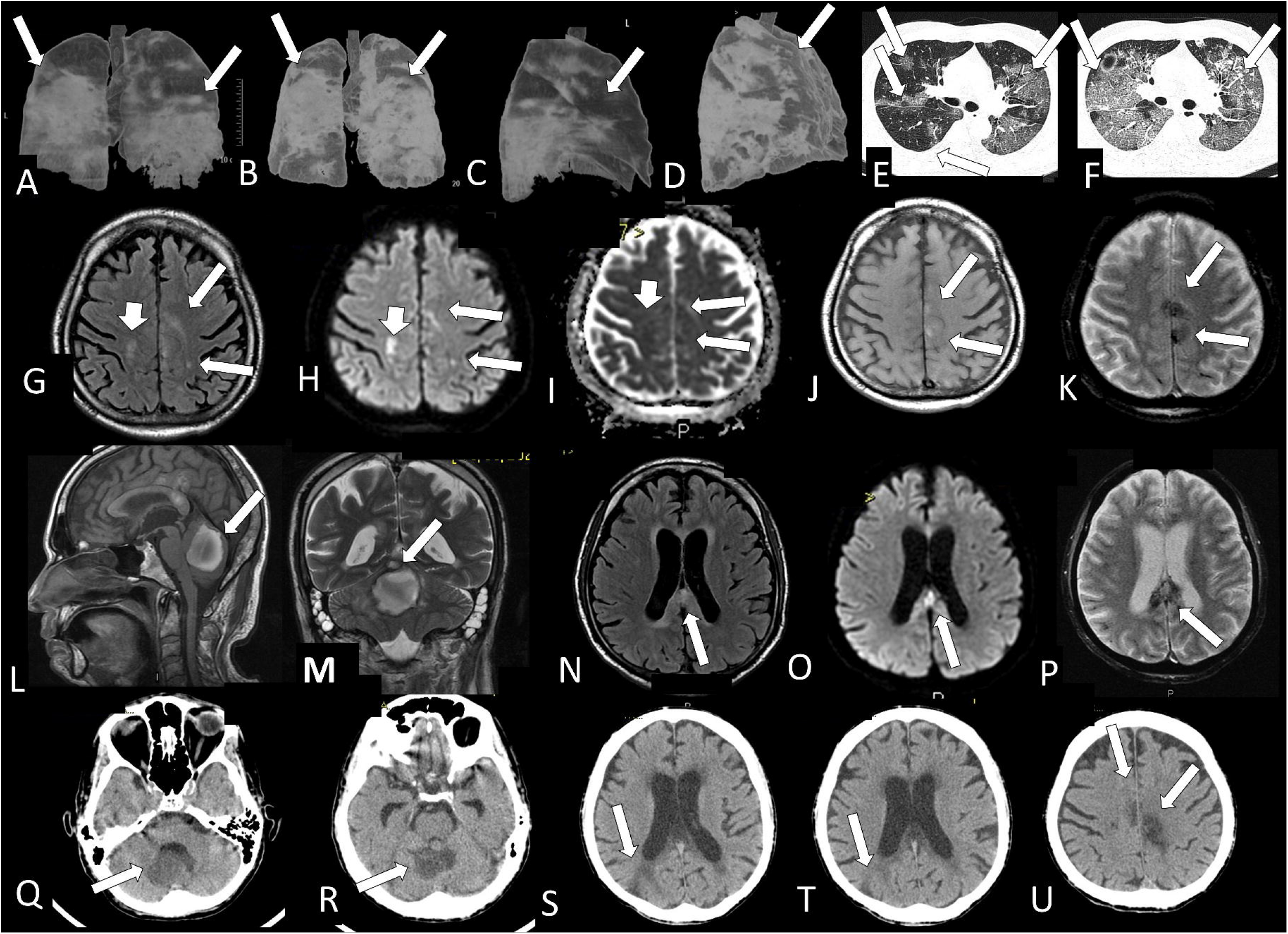
Case 31: A male in his 60’s year-old with confirmed COVID-19 patient admitted to the ICU with fever, cough, dyspnea which needed a mechanical ventilator; had sepsis, stroke and in coma and with a history of diabetes mellitus, systemic arterial hypertension and chronic renal failure. On admission, his chest CT showed on 3D reconstruction posterior view (arrows, A) and lateral view (arrow, C), and on lung window axial CT slices (arrows, E) shows areas of alveoellar consolidation and ground glass which occupy more than 50% of the area of the lungs. Three days later (arrows, B), there was fast progression with severe increase of the lung disease (arrows, B, D, F). The brain MRI shows some hyperintense small areas of on FLAIR and in DWI which are hypointense on ADC-Map (restricted diffusion lesions) in the right semioval center and at left superior frontal gyrus. Small areas of microbledding are also observed, being hyperintense on T1WI (J, arrows) and hypointense on T2* (K, arrows) localized on the left superior frontal gyrus and medial part of left precentral gyrus. There is also a large oval hematoma located at the anterior and superior vermis of the cerebellum with predominant component of methemoglobin inside, being hyperintense on sagittal T1 (L, arrow) and on coronal T2WI (M, arrow) with peripheric hypointense ring of hemosiderin. Some small lesions which represents microbleeding (methemoglobin)are also located on the splenium of the corpus callosum hyperintense on FLAIR (N, arrow) and sagittal T1WI (L) and in DWI (O, arrow) hypointense on T2* (P, arrow). The CT (Q-U) without contrast made one month later showed that the lesions described above on previous MRI are hypointense and that a new sequel in the right parietal lobe (S and T, arrow) had appeared.

- 8/35 (22.9%) cases with any kind of bleeding or microbleeding (Figure 1 K-M and P; Figure 2 E, F and G; Figure 4 A, D and G and Figure 6),

**Figure 2.**
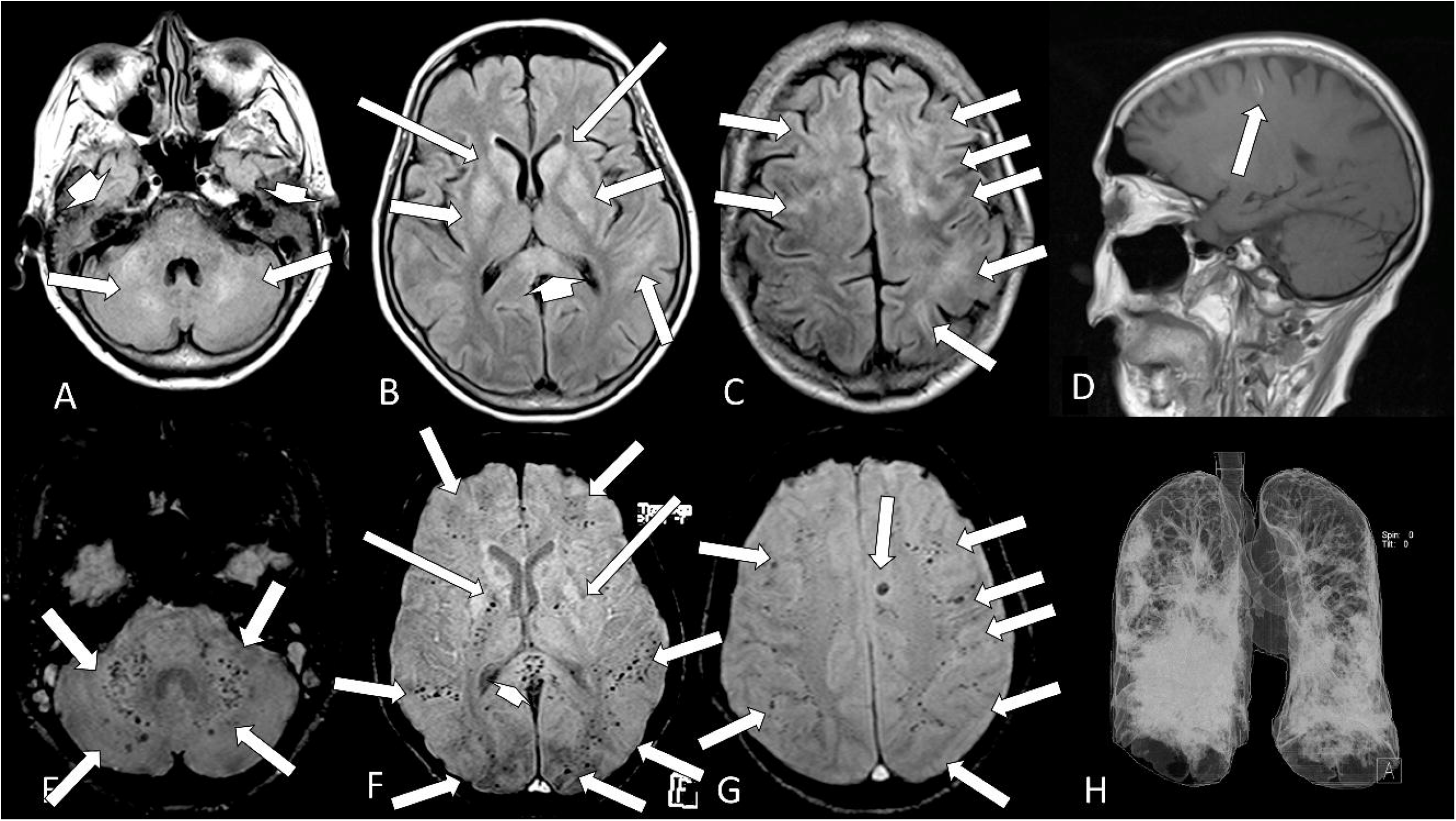
Case 30: A female in her 70’s presented fever, cough, dyspnea, needed a mechanical ventilator, had sepsis which resulted in death. Her brain MRI shows multiple and confluent areas of diffuse hyperintensity on FLAIR (A-C, arrows) and liquid in the mastoid cells (A, short head of arrows). A small linear hyperintense subcortical on sagittal T1 (D, arrow) which represents methemoglobin is observed. There are multiple small dots of micro bleeds at the cerebellum and middle cerebellar peduncle (E, arrows), intern capsule (F, long arrows), splenium of corpus callosum (F, head of arrow) and subcortical white matter (F and G, arrows) of the brain. The 3D chest CT reconstruction shows that there is more than 50% of opacification in both lungs (H).

**Figure 3.**
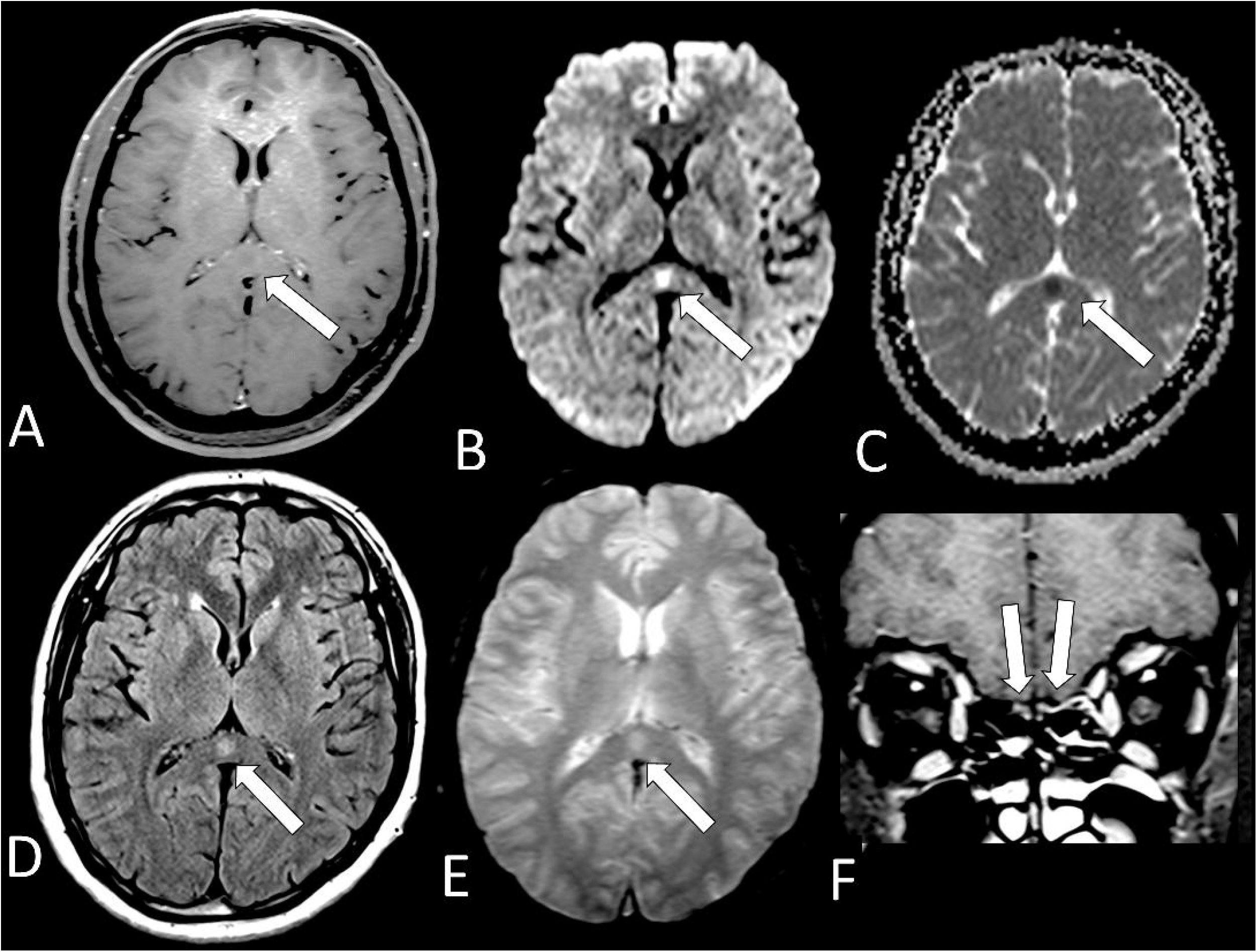
Case 18: Male in his 40’s arrived at the hospital **with** fever, cough, anosmia, headache and sensory alteration in left dimidium, paresthesia in face and fog in visual field. On MRI, there is **a** small round lesion located on the splenium of **the** corpus callosun which could represent a cytotoxic lesion due to a cytokine storm and differential diagnosis is with small acute infartc. It is hypointense on T1 (A, arrow) with restricted diffusion, being hyperintense on DWI (B, arrow) and hypointense on ADC-Map (C, arrow). This lesion is also hyperintense on FLAIR (D, arrow) and T2* (E, arrow) without microbleeding. The olfactory bulbs are hyperintense on coronal fat suppressed T1WI post contrast (F, arrows) ^14^ in comparison with the gray matter of frontal lobes which can represent enhancement, but we can n ot exclude microbleeding^14^.

**Figure 4.**
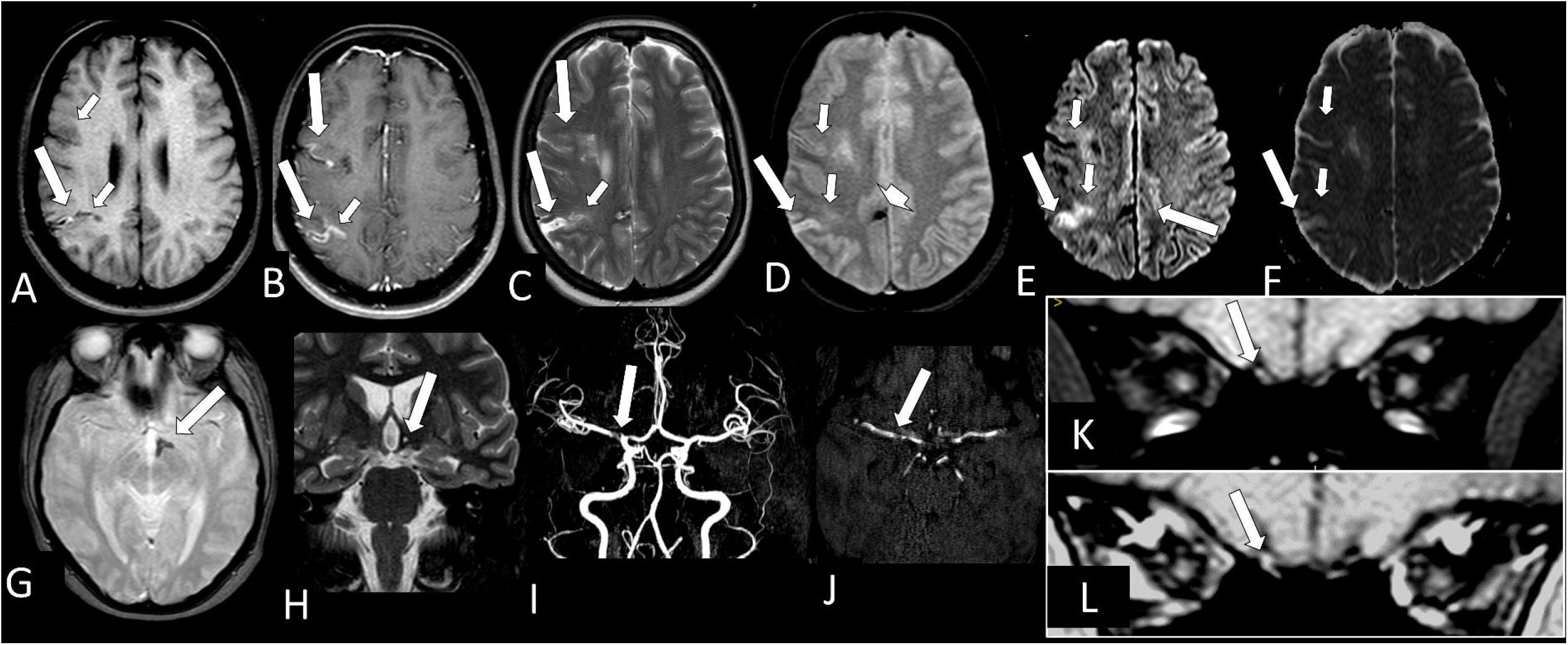
Case 34: A female in her 30’s contracted and recovered from COVID-19 4 months ago and during the acute phase of disease had two strokes and seizures. Her MRI shows a predominantly chronic ischemic stroke, compatible with the patient’s history, located in the cortical and subcortical region of the right middle cerebral artery territory, but specifically in the topography of the right M8 and M10 (A-F). In the right M10, the lesion is a small hyperintense line on T1 (A, long arrow); enhances with contrast (B, long arrow); is hyperintense on T2 and T2*(C and D, long, arrow) and facilitates diffusion, being hyperintense in DWI (E, long arrow) and hyperintense on ADC-Map (E, long arrow), representing T2 shine through effect. Adjacent to this M10 chronic infarct and at M8 lesion, the white matter is hypointense (A, short arrows), with superficial enhanced line on post-contrast T1WI (B, short arrows), and has restricted DWI (E, F, short arrows) being hyperintense on DWI (E, short arrows), and hypointense on ADC-map (F, short arrows), looking like a more recent ischemic stroke, perhaps still representing cytotoxic edema. There are other small areas of bleeding with magnetic susceptibility in T2 * (D, arrow head) located in the right cingulate gyrus and posteriorly and left hypothalamus (G, arrow) which is reduced in volume on coronal T2WI (H, arrow). In magnetic angioMRI, parietal irregularity and narrowing in the right M1 is observed (I and J, arrow) and also in the origin of right anterior artery. Small dot of hyperintensity in the right olfactory bulb (K, arrow) observed on pre contrast fat suppressed T1WI which seems have increased signal with contrast (L, arrow) representing probably methahemoglobine and enhancement.

**Figure 5.**
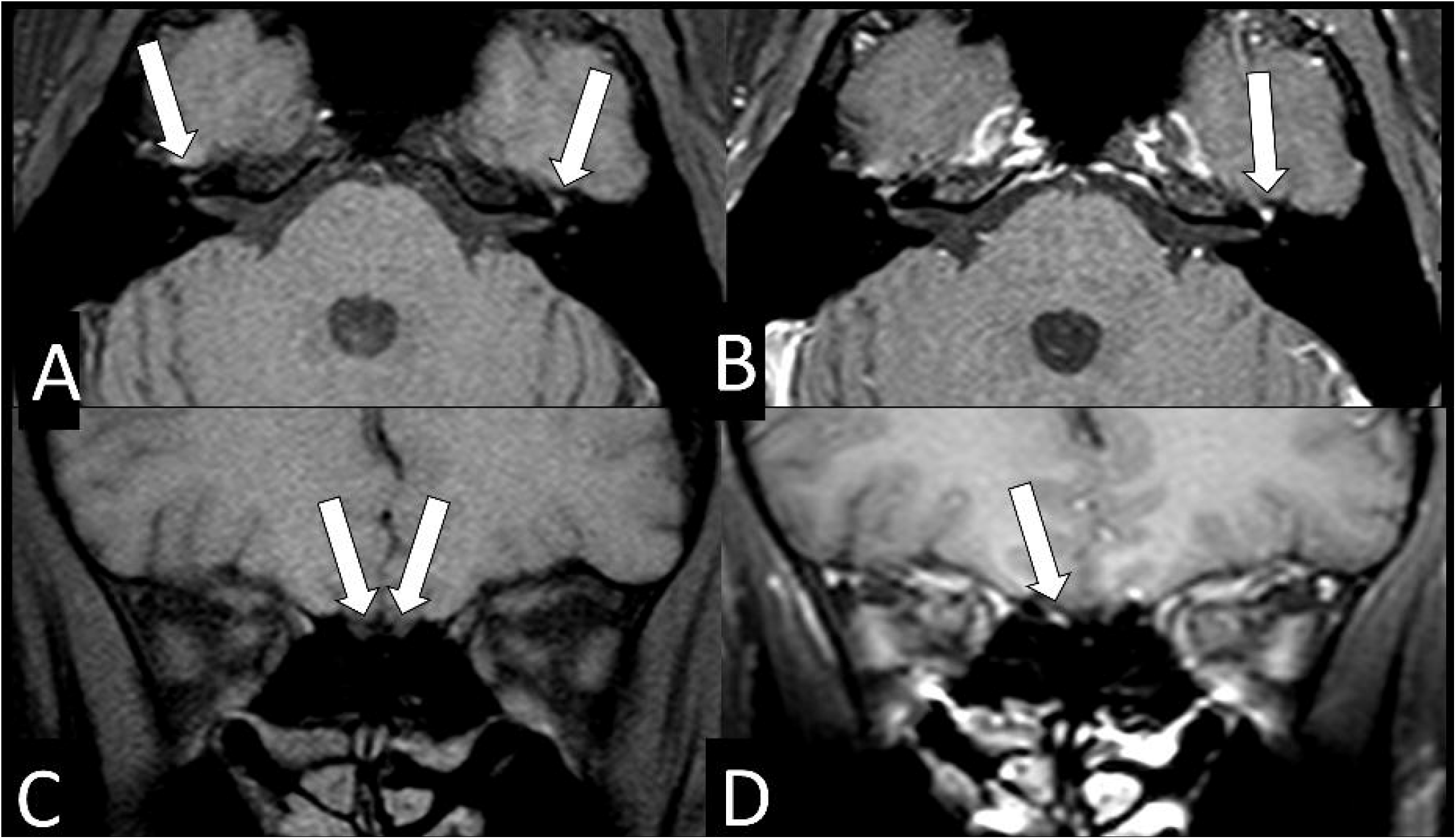
Case 32: A male in his 40’s with COVID-19 developed left peripheric facial palsy around 45 days after the infection. The MRI shows both hypointense geniculate ganglions on axial fat suppressed T1 WI (A, arrows), but the left geniculate ganglion has abnormal stronger enhancement on axial post contrast (B, arrow) fat suppressed T1 WI. The coronal fat suppressed T1 WI shows olfactory bulbs with normal hypointense signal, similar to the gray matter (C, arrows), but the post contrast coronal fat suppressed T1WI shows abnormal enhancement in the right olfactory bulb (D, arrow).

**Figure 6.**
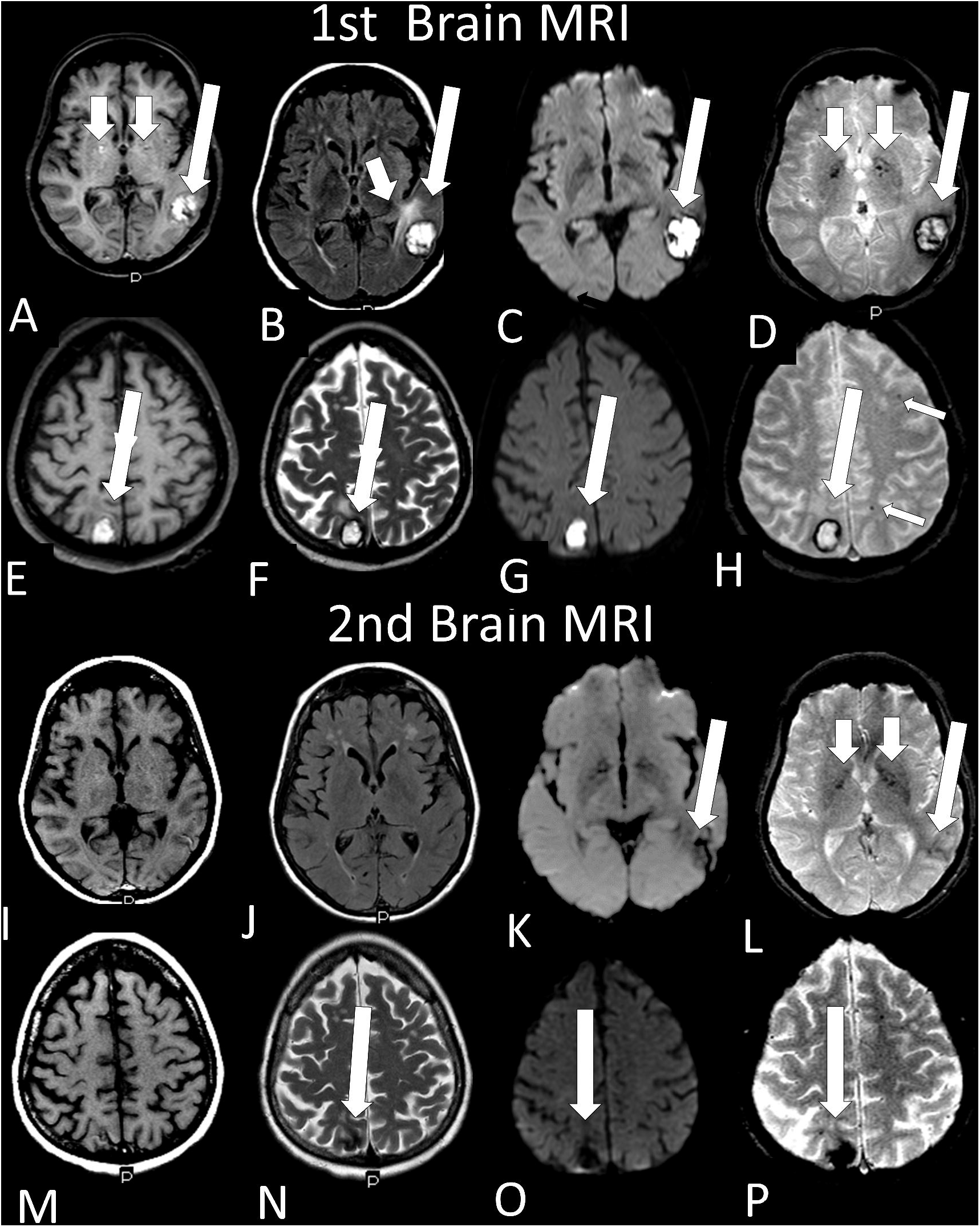
Case 27: Female with COVID 19 in her 60’s has two cortical and cortical and subcortical hematomas on transition between left posterior temporal lobe and occipital lobe (long arrow A-D) and with methemoglobin inside on T1WI (A, long arrow), FLAIR (B, long arrow), DWI (C, long arrow) and T2* (D, long arrow) with hemosiderin on the periphery being hypointense ring. There is vasogenic edema near the left temporal hematoma being hyperintense on FLAIR (B, short arrow). There is also microbleeding in pale globes (A and D, short arrows). Another cortical hematoma in the right parietal lobe (E-H, long arrow, with the same characteristics described above with methemoglobin inside and hemosiderin in the periphery. There are small dots of subcortical microbleeding in the left frontal and parietal lobes (H, short arrows). Two months later there was regression on MRI (I-P) of both hematomas in size, but deposition of hemosiderin remained being hypointense on all sequences [T1, I and M; FLAIR, J; DWI, K and O (arrow); T2, N, (arrow); and T2*, L and P (arrows) and in the lentiform nuclei (L, small arrows).
- 3/35 (8.6%) with restricted diffusion lesions, being; one case with restricted DWI small areas (2.9%) had also associated multiple small areas of microbleeding (Figure 1, H, I); another case (2.9%) had restricted diffusion lesion only in the splenium of the corpus callosun without any microbleeding in the brain (Figure 3 A-E) and with olfactory bulbs injury (Figure 3 F); and one further case (2.9%) of ischemic stroke with 4 months of evolution at the right middle cerebral artery territory (Figura 4 A and B, arrows) with irregularity at the right middle cerebral artery (Figures 4I, 4J, arrows), but with remaining small areas of restricted diffusion (Figure 4 E and F, small arrows), and olfactory bulb injury (Figure 4 K and L, arrows);
- 2/35 (5.7%) with previous old stroke lesion;
- There was a case (2.9%) of left facial nerve palsy with enhacement of geniculate ganglia (Figure 5A and B) and right olfactory bulb/tract injury with enhancement (Figure 5 C and D).

All 12 brain MRIs (100%) which were possible to evaluate the olfactory bulbs (Table 2) had injury in the olfactory bulbs suggestive of enhancement and/or methahemoglobine (Figure 3 F; Figure 4 K, L, Figure 5 C, D and Figure 7 B-D). Six of them (50%) had normal brain MRI and the other six (50%) showed brain MRI abnormalities as well (Table 2).

**Figure 7.**
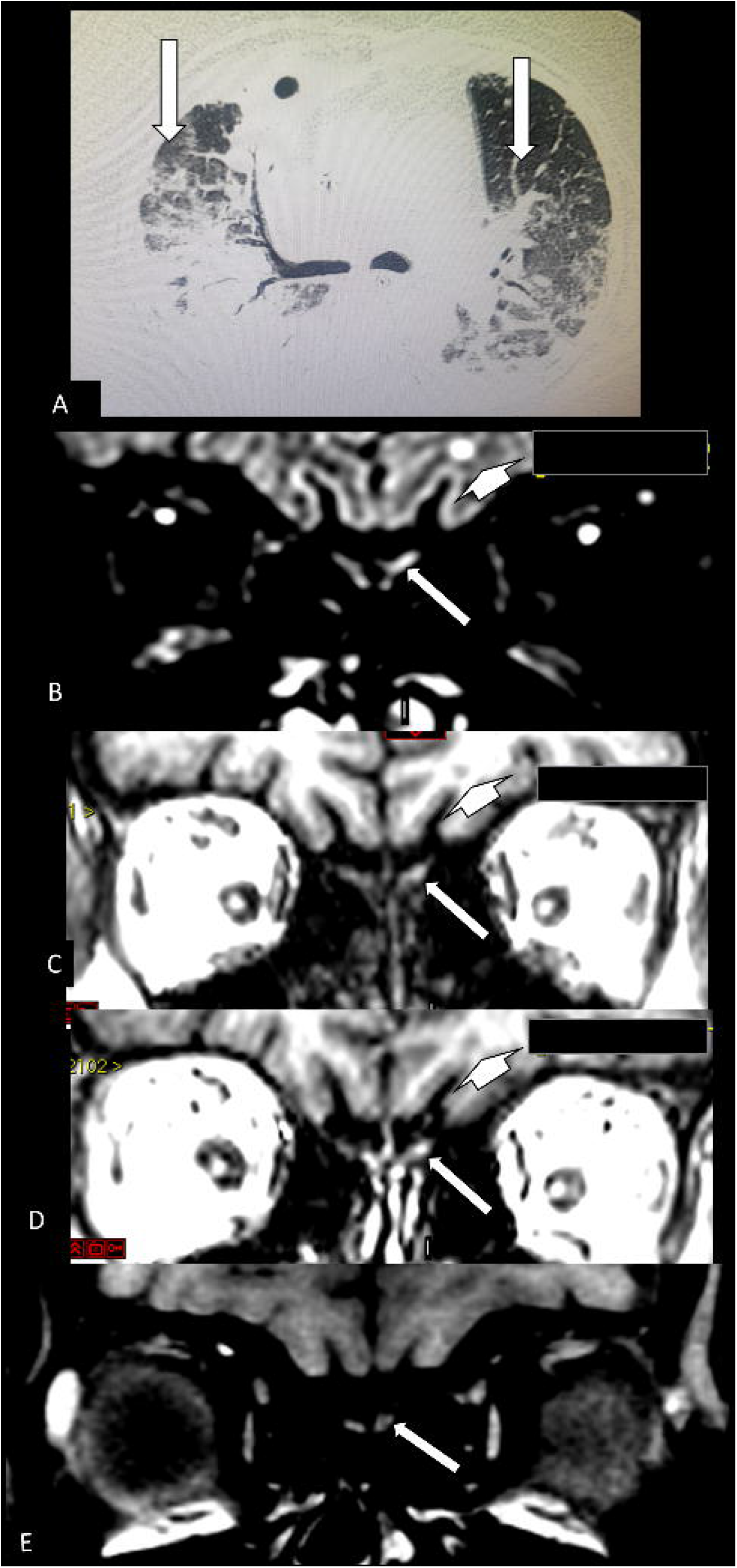
^40^ – The figure 6 continuation of case 27: Her Chest CT (A) shows more than 50% of typical lung opacity of COVID 19 pulmonary injury, bilaterally (arrows). The MRI shows hyperintense lesion on 3D FLAIR (B, arrow)^40^ and also on pre (C, arrow) ^40^ and post contrast SPGR T1 WI (D, arrow) ^40^. This is suggestive of component of probably methemoglobin in this left olfactory bulb lesion which seems be little bigger and asymmetric compared with the apparently normal right olfactory bulb. ^40^ This asymmetry in size is better seen on FLAIR (B). ^40^ There is also a small round hyperintense lesion in the subcortical white matter in the left frontal lobe which is hypointense on T1WI (C, short arrow) and does not enhances on post-contrast T1WI (D, short arrow) being non specific^40^. The MRI done 3 months later shows on fat suppression T1 WI (E) that there is reduction on hyperintensity of left olfactory bulb lesion.

## Discussion

This study shows the impact which only a small percentage of the patients with COVID-19 attended in two private radiologic institutions underwent brain CT or MRI (5.59%), although almost all patients (98,74%) underwent chest CT. This may indicate that patients with COVID-19 rarely have severe neurological complications that require investigation by brain scans, with pulmonary complications being much more frequent and, therefore, chest CT scans are much more performed.

The profile of COVID-19 patients group which needed to undergo brain CT and/or MRI were associated with: more severe disease and admission to ICU, severe lung disease, use of mechanical ventilator, seizure, sepsis and stroke. They had also a statistical tendency for association with chronic renal failure and systemic arterial hypertension.

Despite the COVID-19 patients group which needed to undergo brain CT and/or MRI have severe lung disease and use mechanical ventilator, the complaint of dyspnea was statisticaly less frequent. However, this should be evaluated with caution because dyspnea complaint information was retrieved retrospectively from the medical record, and there may be underreporting.

When anosmia was being analysed in our study, we did not find statistical differences when comparing the radiological group with brain scans to the radiological group with chest CT (without brain scan**s)**. This could be explained because anosmia is a frequent symptom in all COVID-19 patients. Another explanation maybe because a bias arose from this information which was presented as “yes or no” only in 33.2% of the medical records.

Regarding brain imaging findings (without considering olfactory bulbs injury) 40% of the patients had abnormal imaging scans and all of them had vascular brain lesions. Brain haemorrhage lesions (some kind of bleeding or microbleeding) were a more frequent finding in our study and all the patients were 60**+** years old. The second finding was the presence of lesions showing restricted diffusion.

As in previous studies, the most frequent neuroimaging finding was single or multiple T2* punctiform hypointense lesions (microbleeding)^8, 16-19^ (frequently associated with white matter lesions on T2^8, 19^ which sometimes have restricted diffusion^8^) located mainly at the subcortical white matter junction and sometimes at the splenium of the corpus callosum^16-19^.

Several mechanisms for the SARS-CoV-2-related neurological complications are being considered: (a) a direct viral invasion of the haematogenic or retrograde axonal route by olfactory mucosa to olfactory bulbs and to the brain which can lead to intracellular virus accumulation in endothelial cells (endothelitis with thrombotic microangiopathy^16 20^), neurons,^21^ glial cells, macrophages and etc; (b), an indirect process resulting from hypercoagulability-related ^22^; (c) an exaggerated cytokine/^19^immune-mediated response to viral infection causing damage to blood vessel walls or cells in the brain^23^ with (d) ischemia and hypoxic,^16 23^ and (e) treatment complications ^24^or (f) a combination of them^16^.

We had one case in our study of splenium of corpus callosum restricted diffusion lesion without microbleeding. This finding was also described in previous studies ^25, 26^, probably secondary to a cytotoxic lesion^27^ due to a cytokine storm^26^, but but the possibility of acute ischemic injury cannot be ruled out.

The Wuhan study found that severe nervous system disease manifestations were more common in severe infections according to the American Thoracic Society guidelines for community-acquired pneumonia compared with nonsevere infections (45.5% *vs* 30.2%).^28^ Our study found almost the same information in relation to greater pulmonary extension of injury on chest CT which was significantly associated with a radiological group of patients on whom brain scans had been performed because neurological complication.

The primary role of the positive renine angiotensine system is to increase sympathetic nervous system tension, cause vasoconstriction, increase blood pressure, and promote inflammation, fibrosis, and myocardial hypertrophy.^33^ As an organ protector, the whole negative regulatory axis mediated by angiotensine couvert enzyme 2 (ACE2) can antagonize these effects.^33^ However, the ACE2 is known to be a cell receptor for SARS-CoV ^33^ and now confirmed for SARS-Cov2.^34^

A study demonstrates that the ACE2 expression is increased in ischemic brains and vessels of patients with diabetes melitus and exposed to smoking, making them vulnerable to COVID-19 infection.^35^ However, in our study 25% of the patients had diabetes melitus and we did not find any differences between the radiological groups studied. With statistical tendency, systemic arterial hypertension was more frequent in the group submitted to neuroimaging investigation (52.9% vs 22.0%; p = 0.062) maybe for an increased ACE2 expression in this group. It is possible that we did not find statistical differences between both radiological groups (group with brain scans and group with only chest CT) in relations to these variables (diabetes melitus and systemic arterial hypertension) because we are studying patients who needed imaging investigation and for that reason both groups had more severe clinical picture and maybe increased ACE2 expression.

It is worth noting that in COVID-19 the “flu-like syndrome” and mild neurological symptoms^36^ (e.g. headache, anosmia and ageusia) are very common,^37^ but that these symptoms are usually transient, disappearing in around half of the cases within 15 days ^38,39^ and, for this reason, brain imaging (CT or MRI) is not regularly indicated for diagnostic clarification.

COVID-19 patients have presented symptoms of smell dysfunction that is frequently reported in various studies, from 5.1% to 88% of patient^38,39^. However, anosmia in the majority of studies is higher than the central nervous system symptoms which is reported to be around 25%.^28^

However, the occurrence of anosmia or hyposmia is around 16% for other viruses, as for exemple with influenza^37^ and lower than in COVID-19. So, clinical differences can help the clinicians during the co-circulation of influenza and SARS-CoV-2.^37^ Because of this, anosmia is now considered as a criterion for testing COVID-19.

We found only 12 patients with COVID-19 with brain MRI where it was possible to interpret the olfactory bulbs, being: 11 with thin coronal sections pre- and/or post-contrast fat suppression T1 WI in the anterior fossa of the cranium and 1 with pre- and post-contrast SPGR T1WI and FLAIR. All these patients (12/12) had anormalities in olfactory bulbs suggesting microvascular phenomenon, such as microbleeding and/or enhancement by blood-brain barrier breakdown. Of those who had an olfactory bulb injury, 33.33% reported anosmia, 8.33% denied anosmia and in the remaining 58.33% there was no note in the hospital’s Records/Radiology Department about this complaint.

Despite these 12 cases having been investigated for a major neurological complication, only 6 (50%) had other brain lesions and 6 (50%) had normal brain imaging for their age, but all of them had olfactory bulb injury. This finding can be the MRI documentation of olfactory bulbs injury related to the entry of the SARS-CoV-2 from the olfactory mucosa through the cribriform plate into the skull.^14^

The natural history of SARS-CoV-2-induced anosmia has not yet been fully understood. Perhaps if the damage to the olfactory epithelium was more related to an inflammatory process or indirect damage to the neural cells, it would probably explain the cases of evolution with complete recovery. However, there are patients who evolve partial late recovery or even have not yet recovered.

So, our study also suggests and supports the hypothesis that vascular phenomenon could be the main mechanism of COVID-19 in the brain and in the olfactory bulbs injuries^14, 40^ and may also be the main mechanism of anosmia.

Another interesting finding is that despite mortality not being statistically different when comparing the two groups, it was slightly higher among patients investigated by brain neuroimaging (9.1% vs 5.2%).

The limitation of our study is that it is a retrospective study. The variables analyzed were not always noted in the medical records of all patients. Patients underwent brain scans because of major neurological complications and not for anosmia. For this reason, the MRI of most patients did not have an adequate sequence to evaluate the olfactory bulbs. Despite the anterior fossa region being analyzed with coronal fat suppression T1WI can have susceptibility artifacts between the interface with the air, these artifacts are generally well recognized by radiologists and did not hinder the investigation and the analysis.^40^ However, future anatomopathological studies are necessary to confirm our finding.

In conclusion, in COVID-19 patients, the impact of major neurological complication to indicate brain scans was much less frequent than respiratory complication to need chest CT. The profile of COVID-19 patients group which needed to undergo brain scans were statistically associated with the more severe COVID-19 disease, located at ICU, a more severe form of lung disease, use of of mechanical ventilator, complaints of dyspnea (less frequent), seizure, stroke and sepsis. There was a statistical tendency to chronic renal failure and systemic arterial hypertension. Regarding brain imaging findings, less than half of patients had abnormal imaging scans with all of them showing vascular brain injury lesion, being more frequently microbleeding or bleeding, followed by restricted diffusion lesions. All the olfactory bulbs evaluated showed injury by vascular phenomenon, probably methahemoglobine by microbleeding or microthrombus and/or abnormal enhancement by blood-brain barrier breakdown. Only one patient showed left facial nerve palsy with enhacement of geniculate ganglia.

## Data Availability

All the important data (figures of brain MRI and clinical data) of this study is already in the study. However, we obey all ethical rules for the anonymization of patient individual data.
A survey of the cases was carried out to identify the patients with confirmed COVID-19 who underwent chest CT scan and/or brain MRI/CT in two radiology departments in the city of Recife (Brazil): (A) Centro Diagnostico Multimagem and (B) Real Hospital de Beneficencia Portuguesa.
We do not have repositories.

## Acknowledgment

We would like to thank Ms Mirelle Palmeira Lima, MD, Medical Coordinator of the Radiology Sector of the “Real Hospital Português de Beneficência em Pernambuco, Brazil”, for supervising the work of data collection by the authors.

